# Patient Satisfaction and Perception of Community Pharmacy Services in Sidama, Ethiopia: A Mixed-Methods Study Using an Adapted SERVQUAL Framework

**DOI:** 10.1101/2025.08.10.25333113

**Authors:** Daniel Mengistu Kejela, Zerai Hagos

**Affiliations:** Department of Community Development, Leadstar Christian University College, Addis Ababa, Ethiopia

**Keywords:** Patient satisfaction, Community pharmacy, SERVQUAL, Sidama Region, Ethiopia, Healthcare access

## Abstract

**Background:** Community pharmacies are vital for healthcare access in Ethiopia’s Sidama Region, yet patient satisfaction and perceptions of service quality remain understudied. This study evaluates patient satisfaction with pharmaceutical services in urban (Hawassa) and rural (Bensa, Aroresa) settings using a mixed-methods approach and an adapted SERVQUAL framework, incorporating six dimensions: reliability, responsiveness, assurance, empathy, tangibles, and accessibility.

**Methods:** A cross-sectional survey of 400 patients and in-depth interviews with 20 participants were conducted in 2025. Quantitative data were analyzed using descriptive statistics and regression analysis, while qualitative data underwent thematic analysis.

**Results:** Urban patients reported higher satisfaction (M = 4.31, SD = 0.74) than rural patients (M = 3.12, SD = 0.88). Reliability (β = 0.326, p < 0.001) and accessibility (β = 0.244, p < 0.001) were significant predictors of satisfaction. Qualitative findings highlighted challenges such as medicine stockouts, language barriers, and gender-based discomfort, particularly in rural areas. A Sidama-specific SERVQUAL model was developed, emphasizing culturally and linguistically sensitive services.

**Conclusions:** The study reveals urban-rural disparities in pharmacy service satisfaction and underscores the need for targeted interventions to enhance medicine availability, cultural sensitivity, and accessibility in Sidama. The adapted SERVQUAL framework provides a robust tool for evaluating pharmacy services in similar low-resource settings.

## Background

Community pharmacies serve as critical healthcare access points in resource-limited settings, particularly in sub-Saharan Africa, where physician shortages are prevalent [1]. In Ethiopia’s Sidama Region, 62% of residents rely on community pharmacies as their primary source of medical care [2]. However, systemic challenges, including medicine stockouts, limited infrastructure, and communication barriers, undermine service quality, particularly in rural areas [3]. The World Health Organization (WHO) emphasizes patient satisfaction as a key indicator of health system performance and patient-centered care [4]. Ethiopia has implemented reforms like the Auditable Pharmaceutical Transactions and Services (APTS) system to improve pharmacy efficiency [5]. Despite these efforts, disparities persist between urban and rural settings, with rural pharmacies facing frequent stockouts and linguistic barriers [6]. The SERVQUAL model, widely used to assess service quality, has been adapted in low- and middle-income countries (LMICs) to include accessibility and cultural sensitivity [7]. However, its application in Ethiopia, particularly in Sidama, remains limited. This study assesses patient satisfaction and perceptions of community pharmacy services in Sidama using an adapted SERVQUAL framework, incorporating accessibility as a sixth dimension. By combining quantitative and qualitative methods, it aims to provide actionable insights for improving pharmaceutical care in Ethiopia.

## Conceptual Framework

The conceptual framework for this study adapts the SERVQUAL model to the Sidama context, incorporating six dimensions: reliability, responsiveness, assurance, empathy, tangibles, and accessibility. These dimensions are tailored to reflect local cultural, linguistic, and structural factors influencing patient satisfaction in community pharmacies. The framework posits that service quality directly influences patient satisfaction, moderated by contextual factors such as urban/rural residence, insurance coverage, and cultural practices. The adapted framework is visualized below to illustrate the relationships between service quality dimensions, patient satisfaction, and contextual moderators.

## Methods

### Study Design and Setting

A mixed-methods, cross-sectional study was conducted in Sidama National Regional State, Ethiopia, from January to March 2025. Data were collected from public and private community pharmacies in Hawassa City (urban), Bensa Woreda (semi-rural), and Aroresa Woreda (rural), selected purposively for their socioeconomic and geographic diversity.

### Participants

The study included 400 adult patients (≥18 years) who accessed community pharmacy services during the study period. Participants were selected using systematic random sampling from pharmacy logs. For qualitative component, 20 key informants (10 patients, 10 staff) were purposively selected.

### Data Collection

#### Quantitative Data

A structured questionnaire, adapted from the SERVQUAL framework, assessed six dimensions: reliability, responsiveness, assurance, empathy, tangibles, and accessibility. The questionnaire, translated into Amharic and Sidamigna, used a 6-point Likert scale (1 = Strongly Disagree, 6 = Strongly Agree). Socio-demographic data were also collected (Appendix A).

#### Qualitative Data

Semi-structured interviews explored patient and staff experiences, focusing on service delivery, cultural sensitivity, and access barriers. Interviews were audio-recorded with consent and conducted in participants’ preferred languages (Appendix B).

### Data Analysis

#### Quantitative Analysis

Descriptive statistics (means, standard deviations) and multiple linear regression were performed using SPSS v26 to identify predictors of satisfaction. The regression model included SERVQUAL dimensions, urban/rural residence, and insurance coverage as predictors.

#### Qualitative Analysis

Interview transcripts were analyzed thematically using NVivo v12, following a six-step process: familiarization, coding, theme generation, review, definition, and reporting [8]. Themes were triangulated with quantitative findings to ensure robustness.

### Ethical Considerations

Ethical clearance was obtained from Leadstar Christian University College (LCU-2025-014) and the Sidama Regional Health Bureau (SRHB-PH-2025). Informed consent was secured from all participants, with confidentiality and voluntary participation ensured (Appendix C, D).

## Results

### Respondent Characteristics

The sample (N=400) was 54% male and 46% female, with 50% urban and 50% rural residents. Age distribution included 18.5% (18–25 years), 35.3% (26–35 years), 23% (36–45 years), 15.8% (46–60 years), and 7.5% (>60 years). Only 34.8% had health insurance (Table 1).

**Table 1.**

| Variable | Category | Frequency (n) | Percentage (%) |
| --- | --- | --- | --- |
| Gender | Male | 216 | 54.0 |
|  | Female | 184 | 46.0 |
| Age Group | 18–25 | 74 | 18.5 |
|  | 26–35 | 141 | 35.3 |
|  | 36–45 | 92 | 23.0 |
|  | 46–60 | 63 | 15.8 |
|  | 60+ | 30 | 7.5 |
| Residence | Urban | 200 | 50.0 |
|  | Rural | 200 | 50.0 |
| Educational Status | No formal education | 56 | 14.0 |
|  | Primary | 98 | 24.5 |
|  | Secondary | 102 | 25.5 |
|  | Diploma | 84 | 21.0 |
|  | Degree or above | 60 | 15.0 |
| Insurance Coverage | Yes | 139 | 34.8 |
|  | No | 261 | 65.2 |

### uantitative Findings

Mean satisfaction scores were higher in urban areas (M = 4.31, SD = 0.74) than rural areas (M = 3.12, SD = 0.88). Assurance (M = 4.21, SD = 0.82) and reliability (M = 3.67, SD = 0.81) scored highest overall, while accessibility (M = 3.12, SD = 1.02) scored lowest (Table 2). Regression analysis identified reliability (β = 0.326, p < 0.001) and accessibility (β = 0.244, p < 0.001) as the strongest predictors of satisfaction, followed by assurance (β = 0.173, p = 0.001) and empathy (β = 0.087, p = 0.027). Urban residence (β = 0.119, p = 0.011) and insurance coverage (β = 0.128, p = 0.007) also significantly predicted satisfaction (Table 3).

**Table 2.**

| Dimension | Urban (M ± SD) | Rural (M ± SD) | Total (M ± SD) |
| --- | --- | --- | --- |
| Reliability | 4.09 ± 0.74 | 3.25 ± 0.83 | 3.67 ± 0.81 |
| Responsiveness | 3.89 ± 0.85 | 2.95 ± 0.88 | 3.42 ± 0.89 |
| Assurance | 4.55 ± 0.69 | 3.88 ± 0.88 | 4.21 ± 0.82 |
| Empathy | 3.92 ± 0.94 | 2.84 ± 0.91 | 3.38 ± 0.97 |
| Tangibles | 4.12 ± 0.72 | 3.36 ± 0.81 | 3.74 ± 0.77 |
| Accessibility | 3.74 ± 0.91 | 2.62 ± 0.94 | 3.12 ± 1.02 |
| Overall Satisfaction | 4.31 ± 0.74 | 3.12 ± 0.88 | 3.72 ± 0.87 |

| Predictor | B | SE B | Beta (β) | p-value |
| --- | --- | --- | --- | --- |
| Constant | 1.142 | 0.312 | — | < .001 |
| Reliability | 0.365 | 0.063 | .326 | < .001 |
| Accessibility | 0.297 | 0.054 | .244 | < .001 |
| Assurance | 0.211 | 0.061 | .173 | .001 |
| Empathy | 0.109 | 0.049 | .087 | .027 |
| Responsiveness | 0.095 | 0.051 | .072 | .063 |
| Urban Residence | 0.201 | 0.079 | .119 | .011 |
| Insurance Coverage | 0.184 | 0.068 | .128 | .007 |

**Table 3.**

| Theme | Sub-theme | Example Quote |
| --- | --- | --- |
| Stockouts | Unavailability in rural areas | “I came three times and still didn’t get the antibiotic.” — Male, Aroresa |
| Gender Sensitivity | Discomfort with male staff | “I whisper when I ask for family planning.” — Female, Bensa |
| Accessibility | Travel time/distance | “It takes me two hours to reach the pharmacy.” — Female, Aroresa |
| Cultural Practices | Use of traditional medicine | “I use leaves for stomach problems; I don’t always tell the pharmacist.” — Male |
| Language Barriers | Lack of Sidamigna-speaking staff | “Sometimes I don’t understand what they are saying.” — Elder, rural respondent |
| Suggestions for Improvement | Mobile services, female staff, privacy | “If they came to the villages or had a woman to talk to, it would be better.” |

### Qualitative Findings

Five key themes emerged from interviews: stockouts, gender sensitivity, accessibility, cultural practices, and language barriers (Table 4). Rural patients frequently reported medicine unavailability, with one respondent stating, “I came three times and still didn’t get the antibiotic” (Male, Aroresa). Women expressed discomfort discussing reproductive health with male pharmacists, particularly in rural settings. Accessibility challenges included long travel times, with a female respondent from Aroresa noting, “It takes me two hours to reach the pharmacy.” Language barriers were prominent, with elderly rural patients struggling to understand Amharic-speaking staff. Suggestions included mobile pharmacy services, female staff, and enhanced privacy measures.

**Table 4.**

| Dimension | Standard Definition | Sidama-Specific Adaptation |
| --- | --- | --- |
| Reliability | Ability to perform the promised service dependably and accurately. | Consistent medicine availability, accurate dispensing, and adherence to return visit schedules. |
| Responsiveness | Willingness to help customers and provide prompt service. | Staff attentiveness to patient needs, reduced wait times, and willingness to explain and clarify in local language. |
| Assurance | Knowledge and courtesy of employees and their ability to inspire trust and confidence. | Use of name badges, professional dress, respectful tone, and ability to explain medicine clearly—especially to elders. |
| Empathy | Caring, individualized attention to customers. | Gender-matched counseling when possible, respect for traditional beliefs, and sensitivity to women’s privacy concerns. |
| Tangibles | Physical facilities, equipment, and appearance of personnel. | Clean, culturally appropriate pharmacy settings with seating, storage (refrigeration), and privacy spaces. |
| Accessibility | Physical, financial, and informational ease of access to services. | Distance to facility, availability of medicine during holidays or fasting periods, affordability, and language match. |

**Figure 1.**
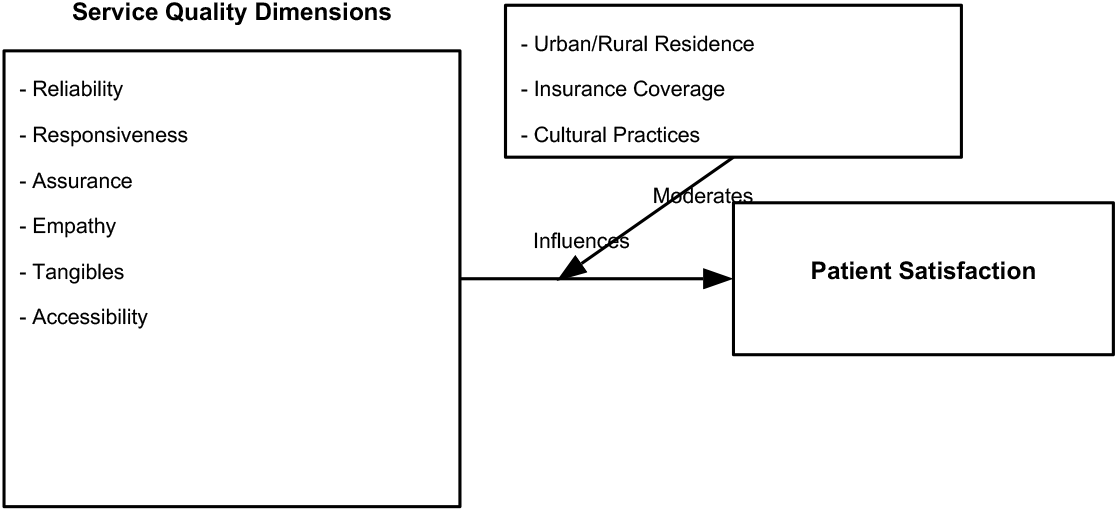
Comparison of Mean SERVQUAL Dimension Scores in Urban and Rural Community Pharmacies, Sidama Region (2025).

**Figure 2.**
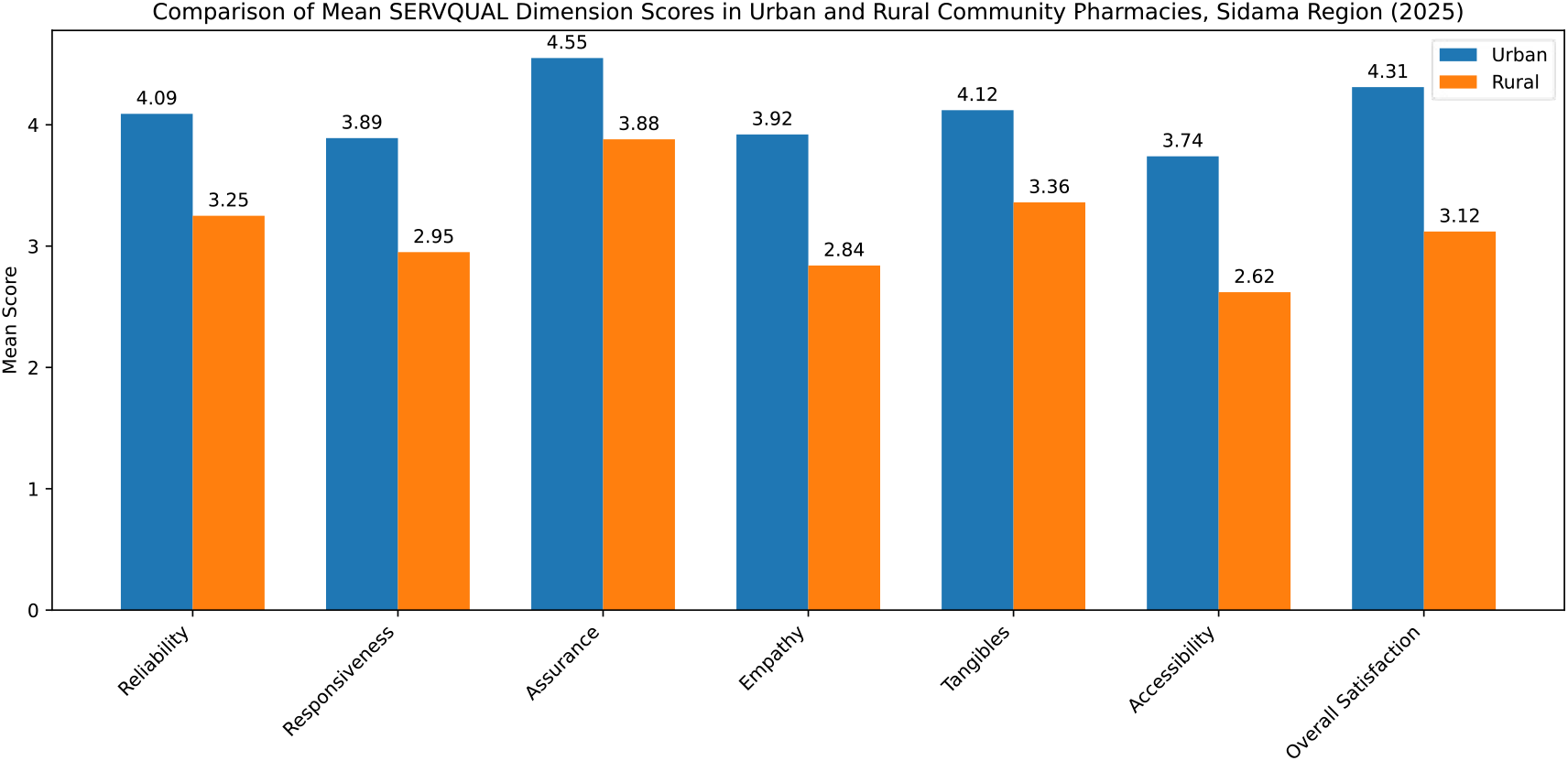
Adapted SERVQUAL Conceptual Framework for Community Pharmacy Services in Sidama, Ethiopia (2025).

## Discussion

This study highlights significant urban-rural disparities in patient satisfaction with community pharmacy services in Sidama, consistent with prior research in Ethiopia [9, 10]. Higher urban satisfaction scores reflect better infrastructure, staffing, and medicine availability, aligning with findings from Addis Ababa [11]. Reliability and accessibility as key predictors underscore the importance of consistent medicine supply and geographic access, particularly in rural areas where stockouts and travel barriers are prevalent [12]. Qualitative findings emphasize cultural and gender-specific challenges. Language barriers, particularly the lack of Sidamigna-speaking staff, align with studies in multilingual African settings [13]. Gender-based discomfort, especially among rural women, highlights the need for gender-sensitive care, as noted in Uganda [14]. The integration of traditional medicine use, often undisclosed to pharmacists, suggests a need for culturally responsive counseling [15]. The adapted SERVQUAL model, incorporating accessibility, proved effective in capturing Sidama-specific service quality nuances. This aligns with adaptations in other LMICs, where accessibility is critical [7]. The proposed Sidama-specific SERVQUAL framework (Table 5) offers a practical tool for evaluating and improving pharmacy services in similar contexts. Limitations: The cross-sectional design limits causal inference, and purposive sampling may reduce generalizability. The small number of qualitative interviews may not fully capture all perspectives. Future studies should include longitudinal designs and broader samples, including non-users of pharmacy services.

## Conclusions and Recommendations

This study provides evidence of urban-rural disparities in pharmacy service satisfaction in Sidama, driven by differences in reliability, accessibility, and cultural sensitivity. To enhance pharmaceutical care, we recommend:

1. Strengthen Medicine Supply Chains: Address stockouts through improved coordination with the Ethiopian Pharmaceutical Supply Service (EPSS).
2. Enhance Cultural Competence: Train pharmacists in Sidamigna and cultural sensitivity to improve communication and trust.
3. Promote Gender-Sensitive Services: Increase female pharmacy staff and private counseling spaces, particularly in rural areas.
4. Expand Accessibility: Pilot mobile pharmacy services and extend operating hours to accommodate rural patients.
5. Adopt the Sidama-Specific SERVQUAL Framework: Use this model for ongoing service quality assessments and policy development.

These interventions can support Ethiopia’s goal of equitable universal health coverage by improving patient-centered pharmaceutical care in Sidama and similar regions.

## Data Availability

https://github.com/HAWASSA2016

